# Phosphatidylethanolamines are the main lipid class altered in red blood cells from patients with VPS13A disease/chorea-acanthocytosis

**DOI:** 10.1101/2024.09.01.24312543

**Authors:** Kevin Peikert, Adrian Spranger, Gabriel Miltenberger-Miltenyi, Hannes Glaß, Björn Falkenburger, Christian Klose, Donatienne Tyteca, Andreas Hermann

**Author notes:** **Corresponding Author:** Prof. Andreas Hermann, MD, PhD; Translational Neurodegeneration Section “Albrecht Kossel”; Department of Neurology; University Medical Center Rostock; Gehlsheimer Straße 20, 18147 Rostock, Germany.

## Abstract

**Background:** VPS13A disease (chorea-acanthocytosis) is an ultra-rare disorder caused by loss of function mutations in VPS13A characterized by striatal degeneration and by red blood cell (RBC) acanthocytosis. VPS13A is a bridge-like protein mediating bulk lipid transfer at membrane contact sites.

**Objectives:** To assess the lipid composition of patient-derived RBCs.

**Methods:** RBCs collected from 5 VPS13A disease patients and 12 control subjects were analyzed by mass spectrometry (lipidomics).

**Results:** While we found no significant differences on the overall lipid class level, alterations in certain species were detected: phosphatidylethanolamine species with both longer chain length and higher unsaturation were increased in VPS13A disease samples. Specific ceramide, phosphatidylcholine and sphingomyelin species were also altered.

**Conclusions:** The presented alterations of particular lipid species in RBCs in VPS13A disease contribute to 1) the understanding of acanthocyte formation and 2) future biomarker identification. Lipid distribution seems to play a key role in the pathophysiology of VPS13A disease.

## Introduction

VPS13A disease (formerly known as chorea-acanthocytosis) is a neurodegenerative disorder of the young adulthood and an important differential of Huntington’s disease^1^. Together with XK disease (McLeod syndrome) it has been classified as neuroacanthocytosis syndrome as it is characterized by striatal degeneration and the presence of deformed red blood cells (RBCs), referred to as acanthocytes^2^. Typical clinical manifestations include a variety of movement disorders (chorea and dystonia with orofacial predominance, in later stages parkinsonism), epilepsy, behavioral and cognitive impairment as well as peripheral neuro- and myopathy^1^.

The autosomal-recessive condition is caused by biallelic pathogenic variants in the VPS13A gene leading in most cases to a complete loss of the respective protein, VPS13A/chorein^3^. VPS13A belongs to a family of 4 proteins, VPS13A-D, that are all related to neurodegenerative or neurodevelopmental disorders, such as hereditary parkinsonism (VPS13C) or ataxia (VPS13D)^4-6^. Only recently, VPS13A has been assigned to the new protein superfamily of bridge-like lipid transfer proteins (BLTPs)^7,8^. Forming hydrophobic grooves that span between two organellar membranes at membrane contact sites, these proteins mediate direct bulk lipid transfer, most likely selective for phospholipids^9^. VPS13A localizes between the endoplasmic reticulum and mitochondria, lipid droplets or the plasma membrane^9^. At the plasma membrane, it has been shown to form a complex with the putative scramblase XK^10,11^. In support of a pathomechanistic role of altered membrane lipid distribution and supply in VPS13A disease, elevated levels of several sphingo- and phospholipids have been recently found in the striatum of VPS13 patients^12^. Also, in Huntington’s disease, a distinct shift in the sphingolipid profile of the caudate has been reported^13^.

In this exploratory study, we aimed to study the lipid composition of RBCs from VPS13A patients for various reasons: 1) RBCs are – besides neurons – prominently affected by the disease as a high proportion are acanthocytic. 2) RBCs are “products” of a complex rearrangement of membranes and organelles during erythropoiesis^14^ potentially requiring membrane lipid transfer. 3) RBCs are – in contrast to brain tissue – easily obtainable. RBC lipid composition may therefore be an ideal biomarker candidate.

## Methods

Five genetically confirmed VPS13 patients (4 males, 1 female, mean age 45.6, min 32, max 52 years) and 12 healthy controls (9 males, 3 females, mean age 40.7, min 23, max 56 years) were included in this study. Demographic and clinical data of patients are shown in Table S1. The study was approved by the ethics committee at the Technische Universität Dresden (EK45022009, EK78022015). All participants gave written informed consent in accordance with the Declaration of Helsinki.

EDTA blood samples were processed in accordance to the sample preparation guidelines from Lipotype GmbH (Dresden, Germany; see supplementary material). Mass spectrometry-based lipid analysis was performed by Lipotype GmbH as previously described using a QExactive Orbitrap mass spectrometer (Thermo Fisher Scientific, Darmstadt, Germany)^15^. Table S2 shows the list of analyzed lipid classes and the respective structural detail level of the analysis.

Lipid data was analyzed in mol% for better comparability. An occupational threshold was applied to filter lipid (sub)species that were not adequately measured to solidify findings (see supplementary material).

Multiple t-tests on all remaining (sub)species were performed (without assuming a consistent standard deviation). In order to account for multiple testing, the Benjamini Hochberg procedure with a false discovery rate of 5% was used. Analysis was performed at different levels: lipid class level, lipid (sub)species level, structural or functional category level as well as grouped by chain length/double bounds number/OH-groups number. For visualization of the results on the lipid (sub)species level, a Vulcano plot was generated.

## Results

There was no statistically significant difference in age and sex distribution between the two groups analyzed in this study (Table S3).

Untargeted lipidomic analysis was achieved by mass spectrometry by screening for 23 lipid classes. In total, 575 lipid species and subspecies were measured. After application of the occupational threshold, 313 lipid species and subspecies remained for further analysis. Out of the 23 classes, 13 were easily detectable including ceramide (Cer), cholesterol (Chol), hexosylceramide (HexCer), lyso-phosphatidylcholine (LPC), phosphatidylcholine (PC), phosphatidylethanolamine (PE), phosphatidylcholine-ether (PE O-), phosphatidylinositol (PI), phosphatidylserine (PS) and sphingomyelin (SM), in agreement with previous reports on RBCs ^16^. Moreover, we were able to detect additional species such as lyso-phosphatidylethanolamine (LPE), phosphatidate (PA) and phosphatidylcholine-ether (PC O-).

On the lipid class level, no relevant differences between healthy controls and VPS13A patients could be detected (Fig 1A). The most distinct difference was seen in the PI class with less mol% in the disease group, however, this did not reach statistical significance (p=0.07, q=0.51). Also, after grouping the lipids into structural (glycerophospholipids, sphingolipids, and sterols) or functional categories (lyso vs. membrane lipids), analysis did not reveal striking differences (Fig 1 B, C), as was the case for the number of double bounds and hydroxyl groups of the lipids (Fig 1 D, E). Likewise, while there was a tendency of decrease of lipids with medium length fatty acid chains (30-36 C-atoms) and of increase of lipids with very long chains (39-44 C-atoms) in the patient group compared to control, none of these differences reached statistical significance (Fig 1 F).

**Fig 1.**
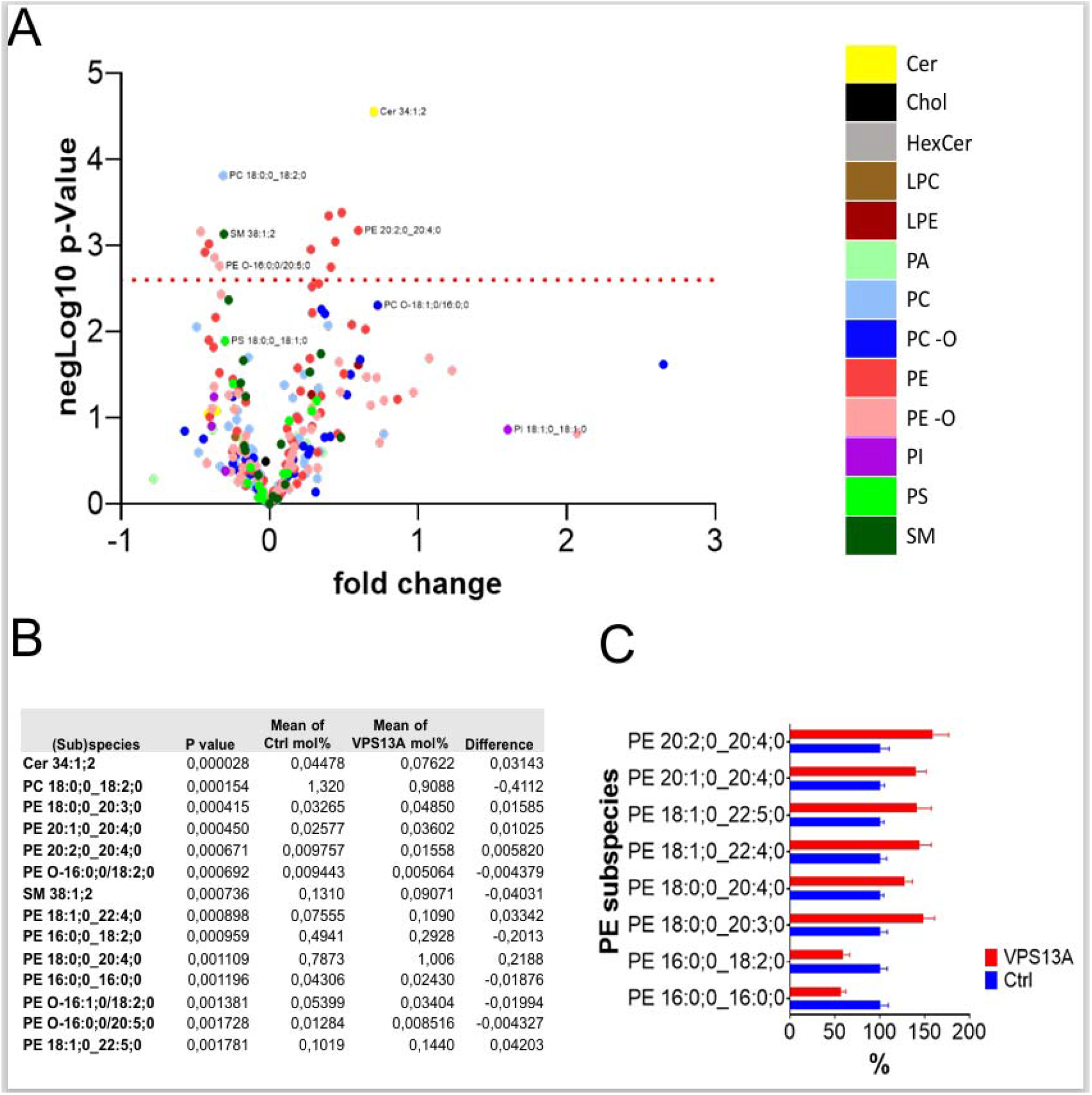
Lipidomics analysis on lipid class (A), structural category (B), functional category (C) level. Structural analysis of lipids in respect to number of double bounds (D), fatty acid chain length (E), and number of hydroxyl groups (F). *Ctrl* controls, *VPS13A VPS13A* patients, *Cer Ceramide, Chol* Cholesterol, *HexCer* Hexosylceramide, *LPC* lyso-Phosphatidylcholine, *LPE* lyso-Phosphatidylethanolamine, *PA* Phosphatidate, *PC (O-)* Phosphatidylcholine (-ether), *PE (O-)* Phosphatidylethanolamine (-ether), *PI* Phosphatidylinositol, *PS* Phosphatidylserine, *SM* Sphingomyelin, *GPL* glycerophospholipids, *SL* sphingolipids, *ST* sterols, *LYS* lyso lipids, *MEM* membrane lipids.

On the lipid species and subspecies level, however, distinct differences with small effect size (fold change) were observed. The increase of Cer34:1;2 was the most significant finding (Fig 2 A, B). Most of the (sub)species with significant change after Benjamini Hochberg procedure belonged to the PE or PE O-classes. Within the PEs, a shift in the fatty chain lengths became obvious: PE subspecies with longer fatty acid chains tended to be increased, species with shorter chains to be decreased (Fig. 2 C). This is in line with the (nonsignificant) shift that has been observed in the overall chain length analysis (Fig. 1 F). In addition, decrease of single PC and SM species were also detected.

**Fig 2.**
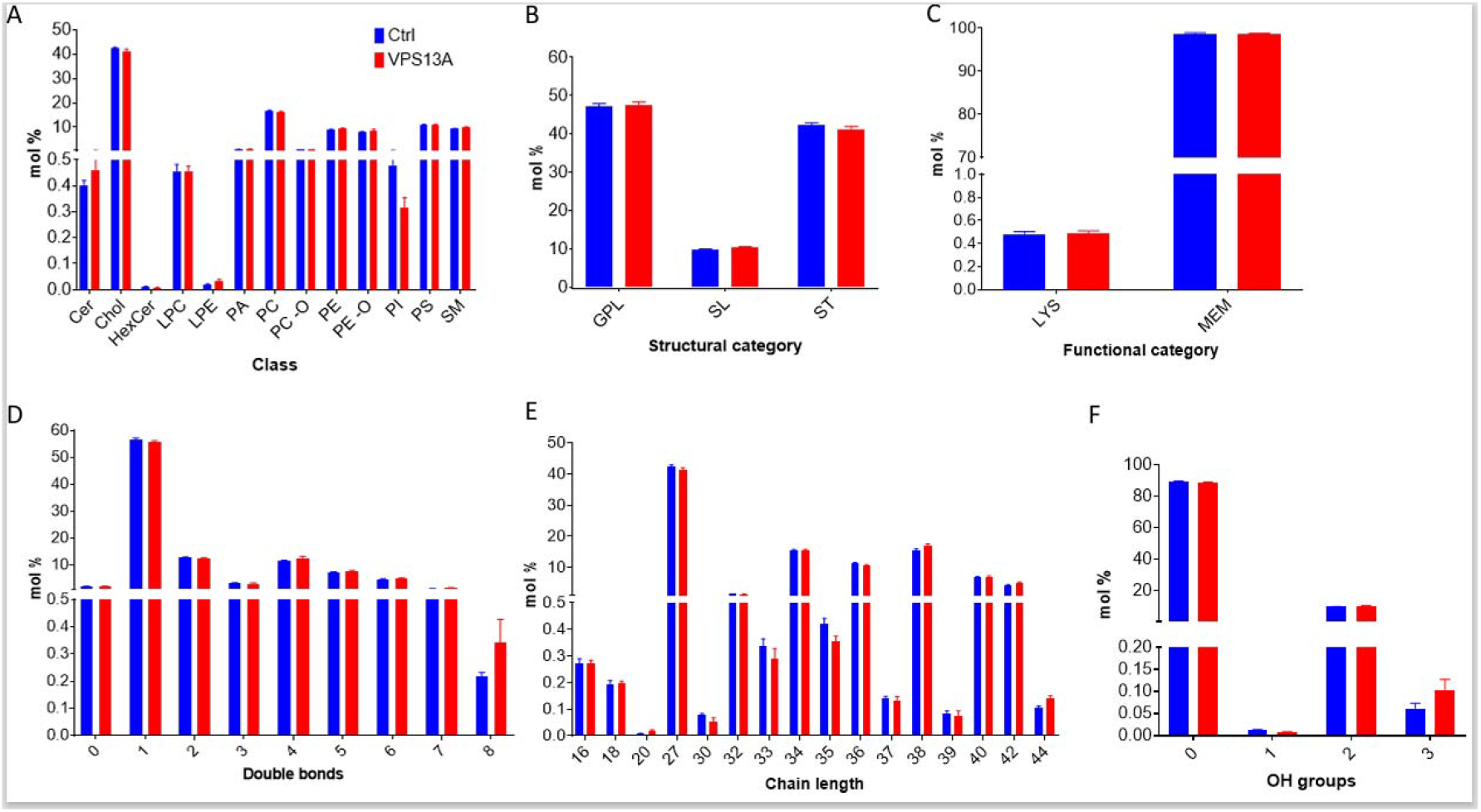
Lipidomics analysis on species and subspecies level. Volcano blot showing all analysed (sub)species (A). The horizonal dotted line represents the threshold of statistical significance after Benjamini Hochberg procedure with a false discovery rate of 5%. (B) shows all significantly different (sub)species between healthy control and VPS13A disease groups. (C) Relative change of Phosphatidylethanolamine (PE) subspecies in VPS13A disease sorted by fatty acid chain length and double bond number; results are expressed as percentage of controls (healthy control values were set 100%). *Ctrl* controls, *VPS13A* VPS13A patients, C*er* Ceramide, *Chol* Cholesterol, *HexCer* Hexosylceramide, *LPC* lyso-Phosphatidylcholine, *LPE* lyso-Phosphatidylethanolamine, *PA* Phosphatidate, *PC (O-)* Phosphatidylcholine (-ether), *PE (O-)* Phosphatidylethanolamine (-ether), *PI* Phosphatidylinositol, *PS* Phosphatidylserine, *SM* Sphingomyelin

## Discussion

VPS13A disease has recently become paradigmatic for a new pathophysiological concept in neurodegeneration: disturbed bulk lipid transfer at membrane contact sites^2,9,17^. VPS13A is a bridge-like protein enabling direct bulk lipid transfer between intracellular membranes. It seems that disturbances of this process are central for VPS13A and related diseases although the exact role of bulk lipid transfer in neuronal and other mainly affected cells such as RBCs is subject of further research.

Based on these recent molecular developments in the field, we studied for the first time the lipid composition of RBCs from VPS13A patients using state-of-the-art lipidomics analysis. RBCs are clearly affected by the disease as acanthocytosis is a core feature and VPS13A deficient RBCs reflect at least partly the pathophysiology of the disease^1,18-20^. Therefore, RBCs may be an easily accessible surrogate and biomarker for pathology of the nervous system. Of note, there is significant evidence that lipid dysmetabolism is causal for the development of acanthocytosis. First, hypo-/abetalipoproteinemia leads to the appearance of acanthocytes ^21,22^. Second, in liver failure where acanthocytosis is observed, irregularities in lipid metabolism, particularly an excess of chol, have been associated with the deformation of RBCs^23^. On the other side, lipid analysis in the “pre-lipidomics era” has not revealed consistent differences in RBCs from neuroacanthocytosis patients^18^.

In line with that, our study did not show a generalized major disturbance of lipid classes, but revealed interesting distinct changes at the lipid species and subspecies level, including PE (O-) and Cer, but also single PC, and SM (sub)species.

PE play a central role in autophagosome formation and is a regulator of autophagy^24^, a process which has been shown to be impaired in VPS13A disease, as reflected by the presence of membrane remnants^19,20^, and which is essential during erythropoiesis. Moreover, the relative abundance of PE species highly evolves during reticulocyte maturation into RBCs: longer and more unsaturated PE species decrease whereas smaller and more saturated PE species increase (Minetti et al, BioRxiv, https://doi.org/10.1101/2023.06.02.543386). Interestingly, we showed here a reverse “acyl chain remodeling”: The former group increased in VPS13A patients while the latter decreased, suggesting impairment of RBC maturation. Therefore, these findings may be related to acanthocyte formation during erythropoiesis and the pathophysiology of VPS13A disease. Supporting this possibility, Cer, PC and PE species were also shown to be affected in hypobetalipoproteinemia, another acanthocyte-related disease^22^. Interestingly, Csf1, a Vps13-like BLTP, has been shown to transfer PEs to the ER for GPI anchor synthesis^25^.

Also, as PE is a non-bilayer forming lipid, especially with longer chain length^26^, an increase of longer PE species might at least partially explain the morphological alterations of acanthocytes. Moreover, non-bilayer lipids may affect integration of proteins into membranes, their lateral movement and their function^27^.

Cer act as the foundational element for complex sphingolipids and is important in cellular signaling. Accordingly, abnormal Cer levels have been associated with several neurodegenerative conditions^28^.

The rather distinct lipid alterations may be viewed as unexpected considering the function of VPS13A as lipid transfer protein. However, as most of the altered lipids are phospholipids, this finding is consistent with the suspected role of VPS13A as (specifically) phospholipid transferring protein^9^. Also, quantitative analyses might not have captured localized changes in membrane composition without major disturbances of overall lipid content. Furthermore, variations within the RBC population (e.g., acanthocytic vs. non-acanthocytic) could have masked specific alterations in a subgroup of cells. As VPS13A plays a role in lipid transfer between organellar membranes and as mature RBCs lack such organelles, the pathogenic process occurs potentially mainly during erythropoiesis. Therefore, quantitative lipid analysis might reveal more pronounced alterations in RBC precursor cells. Another limitation of this study relates to the low number of patient samples due to the ultra-rarity of the disease which might have resulted in an insufficient statistical power to detect even more sub2tle changes.

The RBC lipidomics data presented are not conclusive with the data derived from post mortem brain tissue^12^. This is partially due to the differences in covered lipid classed by the measurements. However, this may also point to different effects of VPS13A deficiency in RBCs and the brain.

In summary, the alterations of particular lipid species in RBCs in VPS13A disease contributes to the pathophysiological understanding. Further studies need to focus on lipid composition of RBC precursor cells and on potential localized changes in distinct RBC membrane domains.

Also, our results may result in the development of PE species as biologically relevant biomarker for VPS13A disease which is a prerequisite for future clinical studies^29^.

## Data Availability

All data produced in the present study are available upon reasonable request to the authors

## Acknowledgments

We thank the patients and control subjects for participating in this study. We are grateful to Glenn (†) and Ginger Irvine as the founders of the Advocacy for Neuroacanthocytosis Patients (www.naadvocacy.org) and to Susan Wagner and Joy Willard-Williford as representatives of the NA Advocacy USA (www.naadvocacyusa.org). We thank the advocacies for their support and research funding. We also thank Dr. Jenny Leopold (Institute for Medical Physics and Biophysics, Faculty of Medicine, Leipzig University, Germany) for her expert advice.

K.P. is supported by the Rostock Academy of Science (RAS), A.H. by the “Hermann und Lilly Schilling-Stiftung für medizinische Forschung im Stifterverband”. D.T. is Senior Research Associate of Belgian F.R.S.-FNRS.

## Authors’ Roles

KP, HG, AH – study design. KP and AH: patient recruitment, KP, AS, HG – execution. KP, AS, GMM, BF, CK, DT – data analysis. KP, AH: funding acquisition. AH: supervision, project administration; KP, AS, GMM, DT – writing. All authors - editing of final version of the manuscript.

## Financial Disclosures of all authors (for the preceding 12 months)

KP: Received funding from the Rostock Academy of Science (RAS) and the Deutschen Gesellschaft für Parkinson und Bewegungsstörungen e.V. Stock Ownership in medically-related fields.

AH has received personal fees and non-financial support from ITF Pharma, Biogen and Desitin during the conduct of the study outside of the submitted work. He received funding from the Schilling Stiftung für medizinische Forschung im Stifterverband, VDI/BMBF, ESF, target ALS foundation outside the submitted work.

CK is CTO and shareholder of Lipotype GmbH.

AS, GMM, HG, BF, DT have no financial disclosure to declare.

## References

1. Peikert K, Dobson-Stone C, Rampoldi L, et al. VPS13A Disease. GeneReviews® [Internet]. University of Washington, Seattle; 1993; 2002 [updated 2023].

2. Walker RH, Peikert K, Jung HH, Hermann A, Danek A. Neuroacanthocytosis Syndromes: The Clinical Perspective. Contact (Thousand Oaks). 2023;6:25152564231210339. doi:10.1177/25152564231210339

3. Dobson-Stone C, Velayos-Baeza A, Filippone LA, et al. Chorein detection for the diagnosis of chorea-acanthocytosis. Ann Neurol. Aug 2004;56(2):299–302. doi:10.1002/ana.20200

4. Velayos-Baeza A, Vettori A, Copley RR, Dobson-Stone C, Monaco AP. Analysis of the human VPS13 gene family. Genomics. Sep 2004;84(3):536–49. doi:10.1016/j.ygeno.2004.04.012

5. Lesage S, Drouet V, Majounie E, et al. Loss of VPS13C Function in Autosomal-Recessive Parkinsonism Causes Mitochondrial Dysfunction and Increases PINK1/Parkin-Dependent Mitophagy. Am J Hum Genet. Mar 3 2016;98(3):500–513. doi:10.1016/j.ajhg.2016.01.014

6. Seong E, Insolera R, Dulovic M, et al. Mutations in VPS13D lead to a new recessive ataxia with spasticity and mitochondrial defects. Ann Neurol. Jun 2018;83(6):1075–1088. doi:10.1002/ana.25220

7. Braschi B, Bruford EA, Cavanagh AT, Neuman SD, Bashirullah A. The bridge-like lipid transfer protein (BLTP) gene group: introducing new nomenclature based on structural homology indicating shared function. Hum Genomics. Dec 02 2022;16(1):66. doi:10.1186/s40246-022-00439-3

8. Kumar N, Leonzino M, Hancock-Cerutti W, et al. VPS13A and VPS13C are lipid transport proteins differentially localized at ER contact sites. J Cell Biol. Oct 1 2018;217(10):3625–3639. doi:10.1083/jcb.201807019

9. Hanna M, Guillén-Samander A, De Camilli P. RBG Motif Bridge-Like Lipid Transport Proteins: Structure, Functions, and Open Questions. Annu Rev Cell Dev Biol. Jul 05 2023;doi:10.1146/annurev-cellbio-120420-014634

10. Park JS, Neiman AM. XK is a partner for VPS13A: a molecular link between Chorea-Acanthocytosis and McLeod Syndrome. Mol Biol Cell. 10 2020;31(22):2425–2436. doi:10.1091/mbc.E19-08-0439-T

11. Guillén-Samander A, Wu Y, Pineda SS, et al. A partnership between the lipid scramblase XK and the lipid transfer protein VPS13A at the plasma membrane. Proc Natl Acad Sci U S A. Aug 30 2022;119(35):e2205425119. doi:10.1073/pnas.2205425119

12. Miltenberger-Miltenyi G, Jones A, Tetlow AM, et al. Sphingolipid and Phospholipid Levels Are Altered in Human Brain in Chorea-Acanthocytosis. Mov Disord. Aug 2023;38(8):1535–1541. doi:10.1002/mds.29445

13. Phillips GR, Saville JT, Hancock SE, et al. The long and the short of Huntington’s disease: how the sphingolipid profile is shifted in the caudate of advanced clinical cases. Brain Commun. 2022;4(1):fcab303. doi:10.1093/braincomms/fcab303

14. Moras M, Lefevre SD, Ostuni MA. From Erythroblasts to Mature Red Blood Cells: Organelle Clearance in Mammals. Frontiers in physiology. 2017;8:1076. doi:10.3389/fphys.2017.01076

15. Sampaio JL, Gerl MJ, Klose C, et al. Membrane lipidome of an epithelial cell line. Proc Natl Acad Sci U S A. Feb 01 2011;108(5):1903–7. doi:10.1073/pnas.1019267108

16. Leidl K, Liebisch G, Richter D, Schmitz G. Mass spectrometric analysis of lipid species of human circulating blood cells. Biochim Biophys Acta. Oct 2008;1781(10):655–64. doi:10.1016/j.bbalip.2008.07.008

17. Peikert K, Danek A. VPS13 Forum Proceedings: XK, XK-Related and VPS13 Proteins in Membrane Lipid Dynamics. Contact2023.

18. Adjobo-Hermans MJ, Cluitmans JC, Bosman GJ. Neuroacanthocytosis: Observations, Theories and Perspectives on the Origin and Significance of Acanthocytes. Tremor Other Hyperkinet Mov (N Y). 2015;5:328. doi:10.7916/D8VH5N2M

19. Lupo F, Tibaldi E, Matte A, et al. A new molecular link between defective autophagy and erythroid abnormalities in chorea-acanthocytosis. Blood. Dec 22 2016;128(25):2976–2987. doi:10.1182/blood-2016-07-727321

20. Peikert K, Federti E, Matte A, et al. Therapeutic targeting of Lyn kinase to treat chorea-acanthocytosis. Acta Neuropathol Commun. May 2021;9(1):81. doi:10.1186/s40478-021-01181-y

21. Levy E. Insights from human congenital disorders of intestinal lipid metabolism. J Lipid Res. May 2015;56(5):945–62. doi:10.1194/jlr.R052415

22. Cloos AS, Daenen LGM, Maja M, et al. Impaired Cytoskeletal and Membrane Biophysical Properties of Acanthocytes in Hypobetalipoproteinemia - A Case Study. Frontiers in physiology. 2021;12:638027. doi:10.3389/fphys.2021.638027

23. Sharma R, Holman CJ, Brown KE. A thorny matter: Spur cell anemia. Ann Hepatol. 2023;28(1):100771. doi:10.1016/j.aohep.2022.100771

24. Hsu P, Shi Y. Regulation of autophagy by mitochondrial phospholipids in health and diseases. Biochim Biophys Acta Mol Cell Biol Lipids. Jan 2017;1862(1):114–129. doi:10.1016/j.bbalip.2016.08.003

25. Toulmay A, Whittle FB, Yang J, et al. Vps13-like proteins provide phosphatidylethanolamine for GPI anchor synthesis in the ER. J Cell Biol. Mar 07 2022;221(3)doi:10.1083/jcb.202111095

26. de Kruijff B. Lipid polymorphism and biomembrane function. Curr Opin Chem Biol. Dec 1997;1(4):564–9. doi:10.1016/s1367-5931(97)80053-1

27. van den Brink-van der Laan E, Killian JA, de Kruijff B. Nonbilayer lipids affect peripheral and integral membrane proteins via changes in the lateral pressure profile. Biochim Biophys Acta. Nov 03 2004;1666(1-2):275–88. doi:10.1016/j.bbamem.2004.06.010

28. Clausmeyer L, Fröhlich F. Mechanisms of Nonvesicular Ceramide Transport. Contact (Thousand Oaks). 2023;6:25152564231208250. doi:10.1177/25152564231208250

29. Peikert K, Glaß H, Federti E, et al. Targeting Lyn Kinase in Chorea-Acanthocytosis: A Translational Treatment Approach in a Rare Disease. J Pers Med. May 10 2021;11(5)doi:10.3390/jpm11050392

