## Supplement for "Phosphatidylethanolamines are the main lipid class altered in red blood cells from patients with VPS13A disease/chorea-acanthocytosis"

**Supplementary Material**

**Data**

**See Supplementary Data table.**

**Methods**

RBC samples were obtained from full EDTA blood. 300 µL of fresh EDTA blood were transferred to 2 mL eppendorf tube followed by a centrifugation with 2000 g, 10 min. The supernatant was discarded and the whitish buffycoat (the leukocyte interlayer) was removed. The tube was filled up to 1,5 mL with "D-PBS without Mg, Ca" and the pellet was resuspended. 300 µL were transferred to another fresh 2mL eppendorf tube and 1,2 mL of "D-PBS without Mg, Ca" were added. After resuspension, another centrifugation step was performed (2000 g, 10 min) and the supernatant once again discarded. After repetition of this step, the tube was filled up to 1,5 mL with water and the pellet was again resuspend. 750 µL were transferred to fresh 2mL eppendorf tube. **Samples were stored at -80˚C and prior to analysis they were thawed at 4˚C.** **Lipids were extracted from this RBC sample using a two-step chloroform/methanol procedure^1^. Samples were spiked with internal lipid standard mixture containing: cardiolipin 16:1/15:0/15:0/15:0 (CL), ceramide 18:1;2/17:0 (Cer), diacylglycerol 17:0/17:0 (DAG), hexosylceramide 18:1;2/12:0 (HexCer), lyso-phosphatidate 17:0 (LPA), lyso-phosphatidylcholine 12:0 (LPC), lyso-phosphatidylethanolamine 17:1 (LPE), lyso-phosphatidylglycerol 17:1 (LPG), lyso-phosphatidylinositol 17:1 (LPI), lyso-phosphatidylserine 17:1 (LPS), phosphatidate 17:0/17:0 (PA), phosphatidylcholine 17:0/17:0 (PC), phosphatidylethanolamine 17:0/17:0 (PE), phosphatidylglycerol 17:0/17:0 (PG), phosphatidylinositol 16:0/16:0 (PI), phosphatidylserine 17:0/17:0 (PS), cholesterolester 20:0 (CE), sphingomyelin 18:1;2/12:0;0 (SM), triacylglycerol 17:0/17:0/17:0 (TAG) and cholesterol D6 (Chol). After extraction, the organic phase was transferred to an infusion plate and dried in a speed vacuum concentrator. 1st step dry extract was re-suspended in 7.5 mM ammonium acetate in chloroform/methanol/propanol (1:2:4, V:V:V) and 2nd step dry extract in 33% ethanol solution of methylamine in chloroform/methanol (0.003:5:1; V:V:V). All liquid handling steps were performed using Hamilton Robotics STARlet robotic platform with the Anti Droplet Control feature for organic solvents pipetting. Mass spectrometry (MS) data acquisition Samples were analyzed by direct infusion on a QExactive mass spectrometer (Thermo Scientific) equipped with a TriVersa NanoMate ion source (Advion Biosciences). Samples were analyzed in both positive and negative ion modes with a resolution of R m/z=200 =280000 for MS and R m/z=200 =17500 for MSMS experiments, in a single acquisition. Tandem mass spectrometry (MSMS) was triggered by an inclusion list encompassing corresponding MS mass ranges scanned in 1 Da increments^2^. Both MS and MSMS data were combined to monitor CE, DAG and TAG ions as ammonium adducts; PC, PC O-, as acetate adducts; and CL, PA, PE, PE O-, PG, PI and PS as deprotonated anions. MS only was used to monitor LPA, LPE, LPE O-, LPI and LPS as deprotonated anions; Cer, HexCer, SM, LPC and LPC O- as acetate adducts and cholesterol as ammonium adduct of an acetylated derivative^3^.**

**Data were analyzed with in-house developed lipid identification software based on LipidXplorer^4,5^. Data post-processing and normalization were performed using an in-house developed data management system. Only lipid identifications with a signal-to-noise ratio >5, and a signal intensity 5-fold higher than in corresponding blank samples were considered for further data analysis.**

**Lipid data was analyzed in mol% for better comparability. An occupational threshold was applied to filter lipid (sub)species that were not adequately measured to solidify findings. For each lipid (sub)species, the number of samples with results below the detection limit (“N/A”) was counted. A respective (sub)species was only included for further analysis if results above the detection limits were given in at least 6 out of 12 assessed samples in the control group and 3 out of 5 samples in the VPS13A patients group.**

**References**

**1.	Ejsing CS, Sampaio JL, Surendranath V, et al. Global analysis of the yeast lipidome by quantitative shotgun mass spectrometry. *Proc Natl Acad Sci U S A*. Feb 17 2009;106(7):2136-41. doi:10.1073/pnas.0811700106**

**2.	Surma MA, Herzog R, Vasilj A, et al. An automated shotgun lipidomics platform for high throughput, comprehensive, and quantitative analysis of blood plasma intact lipids. *Eur J Lipid Sci Technol.* Oct 2015;117(10):1540-1549. doi:10.1002/ejlt.201500145**

**3.	Liebisch G, Binder M, Schifferer R, Langmann T, Schulz B, Schmitz G. High throughput quantification of cholesterol and cholesteryl ester by electrospray ionization tandem mass spectrometry (ESI-MS/MS). *Biochim Biophys Acta.* Jan 2006;1761(1):121-8. doi:10.1016/j.bbalip.2005.12.007**

**4.	Herzog R, Schwudke D, Schuhmann K, et al. A novel informatics concept for high-throughput shotgun lipidomics based on the molecular fragmentation query language. Genome Biol. 2011;12(1):R8. doi:10.1186/gb-2011-12-1-r8**

**5.	Herzog R, Schuhmann K, Schwudke D, et al. LipidXplorer: a software for consensual cross-platform lipidomics. PLoS One. 2012;7(1):e29851. doi:10.1371/journal.pone.0029851**

**6.	Sampaio JL, Gerl MJ, Klose C, et al. Membrane lipidome of an epithelial cell line. Proc Natl Acad Sci U S A. Feb 01 2011;108(5):1903-7. doi:10.1073/pnas.1019267108**

**Supplementary Tables**

| **ID** | **Sex** | **Age range**  **(years)** | **Main clinical manifestation** | **Disease duration**  **(years) ^1^** | **Chorein Western blot^2^** | **Medications** | **Nutritional lifestyle** |
| --- | --- | --- | --- | --- | --- | --- | --- |
| VPS13A_1 | M | 31-35 | Drug resistant epilepsy, mild chorea, tics, cognitive impairment, peripheral neuropathy, myopathy | 9 | Chorein absent | Lacosamide 600 mg/d  Zonisamide 400 mg/d  PRN: Lorazepam/Midazolam | Varied, well-balanced meals, Vitamin D supplementation |
| VPS13A_2 | M | 41-45 | Epilepsy, feeding dystonia, orofacial dyskinesia, chorea, peripheral neuropathy, myopathy, impulse control disorder | 12 | Chorein absent | Levetiracetam 1,5 g/day, Quetiapine  500 mg/day, Ramipril 2.5 mg/day,  Metoprolol 47.5 mg/day, PRN:  metamizole, ibuprofen, pantoprazole | Varied, well-balanced meals |
| VPS13A_3 | F | 46-50 | Epilepsy, parkinsonism, dystonia, dysarthria peripheral neuropathy, cognitive impairment | 28 | Chorein absent | Levetiracetam 4000 mg/d  Valproate 2000 mg/d  Clobazam 10 mg/d  Zonisamide 200 mg/d | Varied, well-balanced meals, Vitamin D supplementation |
| VPS13A_4 | M | 51-55 | Parkinsonism, dystonia, dysarthria, peripheral neuropathy mild depression, | 13 | Chorein absent | Scopoderm transdermal therapeutic  system/day, PRN: Melperone | Varied, well-balanced meals |
| VPS13A_5 | M | 51-55 | Epilepsy, Parkinsonism, dystonia, dysarthria, dysphagia, peripheral neuropathy, cognitive impairment | 19 | Chorein absent | Lamotrigine 550 mg/day,  Oxcarbazepine 1.5 g/day, Lacosamide  300 mg/day, Levodopa 300 mg/day,  Esomeprazole 40 mg/day | Via PEG tube |

**Table S1** Cohort characteristics. M=Male. F=Female. PEG - Percutaneous endoscopic gastrostomy. PRN – pro re nata. ^1^ Since onset of first symptoms. ^2^ Additionally, all cases have been confirmed by genetic testing.

| **Lipid class** | **Abbreviation** | **Mass spectrometry mode** | **Structural detail level** |
| --- | --- | --- | --- |
| Cholesterol esters | CE | Tandem mass spectrometry | Subspecies |
| Ceramide | Cer | Mass spectrometry | Species |
| Cholesterol | Chol | Mass spectrometry | Species |
| Diacylglycerol | DAG | Tandem mass spectrometry | Subspecies |
| Hexosylceramide | HexCer | Mass spectrometry | Species |
| lyso-Phosphatidate | LPA | Mass spectrometry | Species |
| lyso-Phosphatidylcholine (-ether) | LPC/ LPC O- | Mass spectrometry | Species |
| lyso-Phosphatidylethanolamine (-ether) | LPE/ LPE O- | Mass spectrometry | Species |
| lyso-phosphatidylglycerol | LPG | Mass spectrometry | Species |
| lyso-Phosphatidylinositol | LPI | Mass spectrometry | Species |
| lyso-Phosphatidylserine | LPS | Mass spectrometry | Species |
| Phosphatidate | PA | Tandem mass spectrometry | Subspecies |
| Phosphatidylcholine (-ether) | PC/ PC O- | Tandem mass spectrometry | Subspecies |
| Phosphatidylethanolamine (-ether) | PE/ PE O- | Tandem mass spectrometry | Subspecies |
| Phosphatidylglycerol | PG | Tandem mass spectrometry | Subspecies |
| Phosphatidylinositol | PI | Tandem mass spectrometry | Subspecies |
| Phosphatidylserine | PS | Tandem mass spectrometry | Subspecies |
| Sphingomyelin | SM | Mass spectrometry | Species |
| Triacylglycerol | TAG | Mass spectrometry | Species |

**Table S2** List of analyzed lipid classes, their abbreviation used in this manuscript, mode of mass spectrometry and structural detail level.

| **Cohort** | **VPS13A** | **Controls** | **p value** | **statistic test** |
| --- | --- | --- | --- | --- |
| **Age (years)** | 45.6±8.5 | 40.7±11.6 | 0.4 | Mann Whitney |
| **Sex (M:F)** | 4:1 | 9:3 | 0.8 | Chi square |

**Table S3** Cohort characteristics. Data are presented as mean±SD. M=Male. F=Female.
